# Racial and workplace disparities in seroprevalence of SARS-CoV-2 in Baton Rouge, Louisiana, July 15-31, 2020

**DOI:** 10.1101/2020.08.26.20180968

**Authors:** Amy K. Feehan, Cruz Velasco, Daniel Fort, Jeffrey H. Burton, Eboni Price-Haywood, Peter T. Katzmarzyk, Julia Garcia-Diaz, Leonardo Seoane

**Affiliations:** Ochsner Clinic Foundation, New Orleans, LA, USA (A. K. Feehan, C. Velasco, D. Fort, J. H. Burton, E. Price-Haywood, J. Garcia-Diaz, L. Seoane); The University of Queensland Faculty of Medicine, Ochsner Clinical School, New Orleans, LA, USA (A. K. Feehan, J. Garcia-Diaz, E. Price-Haywood, L. Seoane); Pennington Biomedical Research Center, Louisiana State University, Baton Rouge, LA, USA (P.T. Katzmarzyk); Louisiana State University Health Sciences Center-Shreveport, Shreveport, LA, USA (L. Seoane)

**Keywords:** SARS Virus, COVID-19, Cross-Sectional Studies, Healthcare Disparities, Prevalence, Seroepidemiologic Studies, Baton Rouge, Convalescence

## Abstract

Using paired molecular and antibody testing for SARS-CoV-2 infection, we determined point prevalence and seroprevalence in a municipality in Louisiana, USA during the second phase of reopening. Infections were highly variable by race, work environment, and ZIP code. Census-weighted seroprevalence and point prevalence were 3.6% and 3.0%, respectively.

## Text

Previously we reported results from a seroprevalence study conducted in New Orleans, Louisiana, USA, hit hard early in the COVID-19 pandemic (1). Baton Rouge is a large metropolitan area roughly 80 miles northwest of New Orleans and at the time of this study was in the second phase of reopening following a stay-at-home order. While the seroprevalence in New Orleans (6.9%)(1) was similar to prevalence recorded in Spain (5%), Sao Paulo, Brazil (4.7%) and New York, USA (6.9%) (2,3,4), Baton Rouge had only 3,427 more cases as of August 2, 2020 (17,093 cases) than New Orleans did by May 16, 2020 (13,666 cases) (5). This study was designed to estimate SARS-CoV-2 infections in the greater Baton Rouge area (Ascension, East Baton Rouge, Livingston, and West Baton Rouge Parishes) with additional information on potential workplace exposures.

The protocol was approved by the Ochsner IRB and intended to enroll and test up to 2,500 participants at 13 sites throughout Baton Rouge between July 15-31. Recruitment targeted a representative sample in a method developed by Public Democracy (www.publicdemocracy.io) and described elsewhere (1,6). In contrast to the New Orleans study, where people tested were under a “stay-at-home” order, Baton Rouge was in phase two of reopening. A randomized subset of 500,000 Baton Rouge residents was targeted with digital ads for recruitment. From those, 3,687 volunteers were recruited and re-stratified by census designations: 2,309 were invited, 2,179 were enrolled and completed testing, and 2,138 were included in the final analysis. Thirty-eight subjects were excluded for living in ineligible ZIP codes and 3 withdrew consent (Supplemental Figure 1). All study materials were in English, Spanish, and Vietnamese. Participants were offered free transportation. Research staff verbally consented subjects and electronically documented consent and survey responses. Subjects then had a blood draw and nasopharyngeal (NP) swab performed.

US Food and Drug Administration-Emergency Use Authorization approved tests were used. Real-time reverse transcriptase polymerase chain reaction (PCR) tests of NP swabs were performed on the Abbott m2000 RealTime system. Qualitative immunoglobulin G (IgG) blood tests were performed on the ARCHITECT i2000SR. The IgG test meets criteria described by the CDC to yield high positive predictive value, which was validated by Ochsner Health laboratory and by others (7,8). Study participants with either or both positive tests were assessed as having been infected with SARS-CoV-2. Point estimates and corresponding 95% confidence intervals (CI) for proportions of COVID-19 exposure (PCR+ and/or IgG+ tests), point prevalence (PCR+, IgG-), and seroprevalence (IgG+ tests regardless of PCR test) were estimated for the Baton Rouge area using raw and census-weighted counts. Unadjusted odds ratios with Firth correction were calculated for all variables.

The sample was 63.6% female, 66.9% white, average age of 48.7 years, and average household size of 2.84 people. The census-weighted SARS-CoV-2 infections in the sample is 6.6% (6.0%, raw) with 3.0% positive for active viral shedding without detectable antibody, which translates to 16,536 contagious individuals. By race, seroprevalence was highest (7.5%) in Black subjects which was higher than any other group: White non-Hispanic (1.8%), Asian non-Hispanic (1.7%), Hispanic of any race (1.6%) and Other (2.7%). Sample breakdown and population estimates for adults over 18 years old are provided in Table 1 along with raw and weighted exposure, weighted point prevalence, and weighted seroprevalence.

**Table 1.**
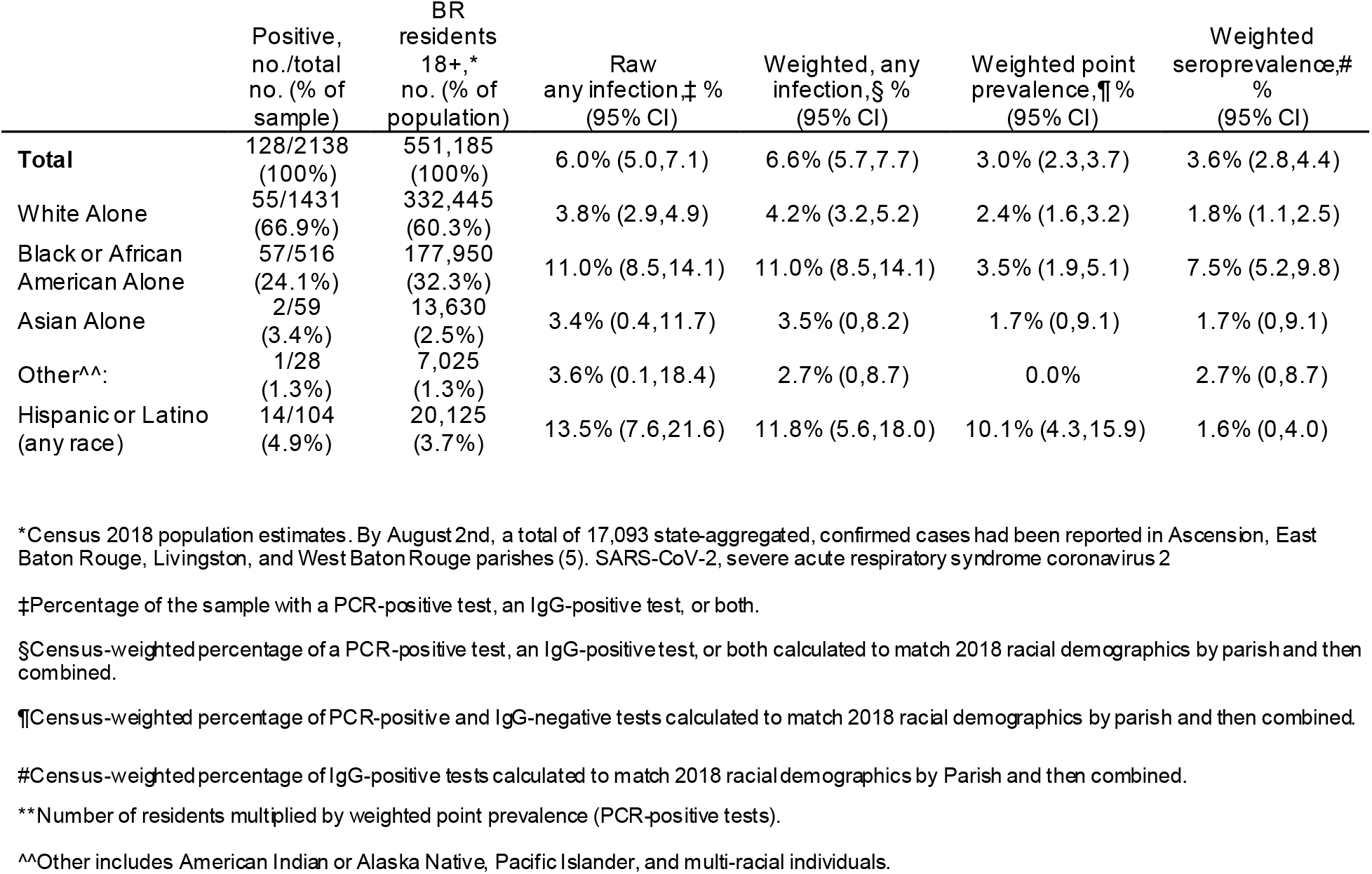
Prevalence of SARS-CoV-2 past and present infections by race and ethnicity across Baton Rouge, LA after phased reopening.

The point prevalence and any SARS-CoV-2 infection are listed and mapped by ZIP codes across the greater Baton Rouge area (Supplemental Figure 2). Point prevalence and all infections were highly variable by ZIP.

Marital status was associated with prevalence (p=0.0005). Single individuals were the most infected (9.3%) compared with married or cohabitating subjects (5.0%) and were 1.9 times more likely to test positive (Figure 1). Work environment was also associated with prevalence (p=0.01) with the lowest prevalence in the group who worked from home (WFH) part time and went in to work part time (3.7%). Those who were mostly going into work had the highest prevalence (8.2%) and were 2.3 times more likely to test positive than those who WFH part time. Infections varied by type of job reported (p=0.01) with the lowest positivity in office workers (3.0%) and increased odds of testing positive in public, delivery, healthcare and other jobs. However, based on seroprevalence, which was also significantly different by job type (p=0.03), healthcare workers and public-facing workers bore the brunt of early infections shown by the higher odds of testing antibody-positive (Figure 1).

**Figure 1.**
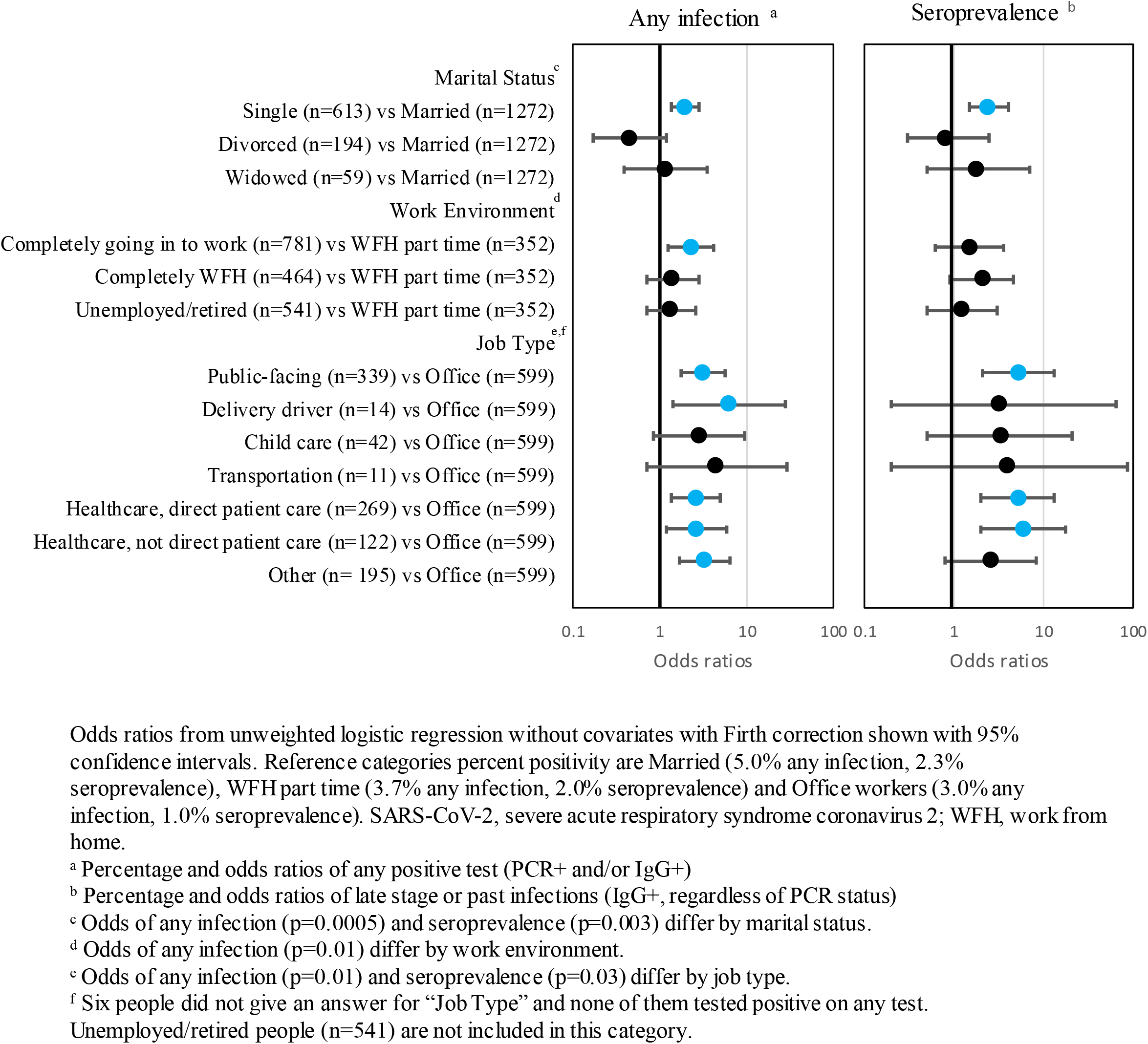
Odds ratios of SARS-Cov-2 infections by marital status, work environment, and job type in Baton Rouge, LA, after phased reopening.

This study found the prevalence of SARS-CoV-2 infection in Baton Rouge to be 6.6%, but was heavily concentrated on new, contagious infections (3.0%). There is a chance that those infected very early on no longer had antibodies. This differed from the New Orleans study performed after extensive lockdowns with 0.9% new infections (1). Some populations had higher rates of infection than others, including Black and Hispanic communities and public-facing workers or those who do not WFH.

## Data Availability

De-identified data is available upon reasonable request to the corresponding author.

## Acknowledgments

The authors would like to especially thank the labs at the Ochsner Medical Center Jefferson Highway Campus for testing and keeping track of research samples; Christy Reeves for liaising with public leaders and sites; Eric Sapp and Dan Nichols from Public Democracy for their recruitment effort; Susan Green, Charlene Ho, Lena Hooper, Patty Kline, and Candace Melancon for clinical site management; Johanna Veal, Lyndsey Buckner-Baiamonte, Ansley Hammons, and Ashley LaRoche for research site management; and countless research coordinators, clinical staff, marketing personnel, medical students, and Epic and IT staff for making site testing possible. The Ochsner Health Market Planning and Analysis team designed the maps in supplemental Figure 1. The authors thank Kathleen McFadden for her thorough editing. We would like to thank the Ochsner Language Services Department for helping to increase inclusivity. We would also like to acknowledge the East Baton Rouge Mayor-President Sharon Weston Broome and Pennington Biomedical Research Center for their collaboration and support of this project.

## Funding

The study was funded by the Baton Rouge Area Foundation, Louisiana COVID-19 Health Equity Task Force, and The Humana Foundation, with additional support from The Blue Cross and Blue Shield of Louisiana Foundation, Healthy Blue, the Huey and Angelina Wilson Foundation, and the Irene W. and C.B. Pennington Foundation.

The funders had no role in study design, data collection and analysis, decision to publish, or preparation of the manuscript.

---

**Supplementary Figure 1.**
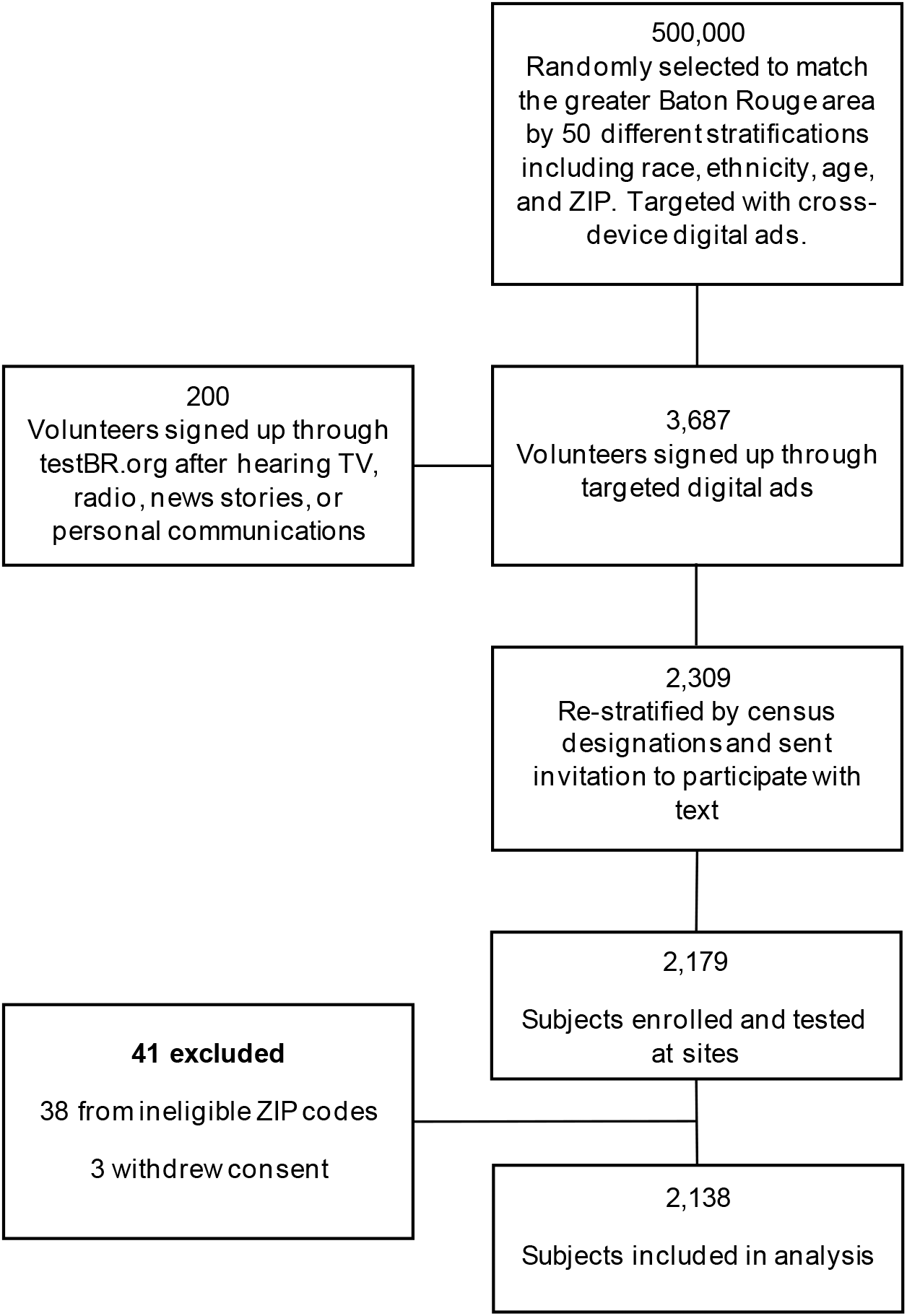
Flow diagram of recruitment to enrollment and analysis.

**Supplementary Figure 2.**
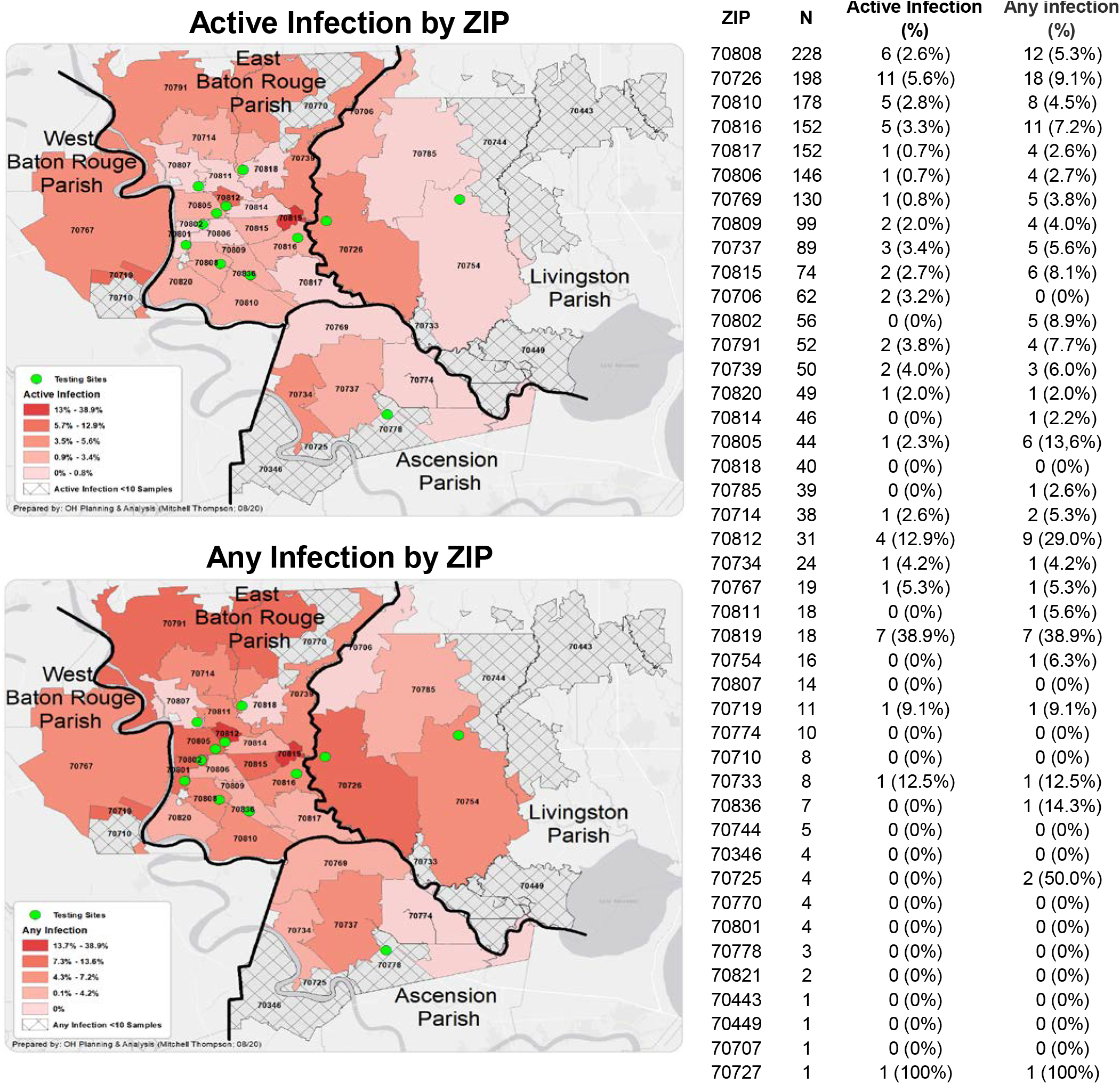
Heatmaps of the Greater Baton Rouge Area (Ascension, East Baton Rouge, West Baton Rouge, and Livingston parishes) showing (top) active infections and (bottom) any infections by ZIP code. Number of participants, positives and percentages are listed by ZIP on the right.

## Notes

### Competing Interest Statement

The authors have declared no competing interest.

### Author Declarations

The Ochsner Health IRB approved this study and can be contacted at or by phone at 504-842-3535.

